# Clinical Retrospective Analysis: Higher Rates of Gene Mutations in the Microsatellite Stable (MSS) Population of Colorectal Cancer

**DOI:** 10.1101/2024.03.25.24304166

**Authors:** Xionglin Liu, Yuan Zhou, Wei Mao, Haiping Pei

## Abstract

With the increasing incidence of colorectal cancer, understanding different subgroups, such as the microsatellite stable (MSS) population, is crucial. Our study aimed to explore the unique characteristics and underlying genetic and epigenetic features of the microsatellite stable (MSS) population within colorectal cancer. Using 325 samples, we compared MSS with other microsatellite statuses on more than 50 clinical indicators. A significantly different overall gene mutation rate was observed in the MSS group compared to the other groups, especially in P53 mutations. No notable changes were found in epigenetics. The results suggest that MSS colorectal cancer cases are more likely to have gene mutations than other subgroups. We also found that the N stage was lower in the MSI-H group. These findings provide crucial insights that can guide future drug development and treatment plans for the MSS population.

## Introduction

Colorectal cancer is one of the most common malignant tumors worldwide, with consistently high rates of incidence and mortality. From an epidemiological perspective, colorectal cancer is one of the leading causes of cancer-related deaths in many developed and developing countries and is closely related to diet, lifestyle, and genetic factors^[1]^.

Approximately 15% of colorectal cancer populations exhibit MSI (microsatellite instability) mutations. MSI primarily results from mutations in the cellular mismatch repair mechanism, leading to deficiencies in mismatch repair proteins (dMMR)^[2]^. The general pathway is as follows: Initial mutations or unexpected events such as promoter methylation led to dMMR, primarily involving four proteins—MLH1, PMS2, MSH2, MSH6^[3]^. This results in genetic instability in the noncoding and microsatellite regions, which in turn causes alterations in the noncoding areas of oncogenes and tumor suppressor genes, affecting normal gene expression^[4]^. Tumors with MSI are significantly associated with Lynch syndrome^[5, 6]^. Due to their higher mutational burden, MSI tumors are capable of generating more neoantigens, making them more likely to be recognized and attacked by the immune system^[7]^. Therefore, MSI-high tumors tend to respond well to immunotherapies, particularly PD-1 inhibitors. Pembrolizumab (Keytruda) is a PD-1 inhibitor that works by blocking the PD-1 protein, effectively lifting the tumor’s suppression of the immune system and allowing the immune system to attack the tumor cells more effectively^[8]^. In 2017, the U.S. FDA approved pembrolizumab for treating MSI-high solid tumors. This was the first time an anticancer drug was approved based on a tumor’s molecular marker rather than its tissue type^[9]^. Previously, patients with high MSI (MSI-H) colorectal cancer typically had a poorer prognosis^[10]^. Currently, with the advent of PD-1 inhibitors and other drugs, they have a better outlook and may respond better to certain immunotherapies^[11]^.

The evolving landscape of colorectal cancer treatment has primarily centered on tumors with microsatellite instability (MSI)^[12]^, yet a significant number of cases remain microsatellite stable (MSS). The MSS subgroup has often been overlooked in research, despite generally presenting with poorer prognoses. Understanding the unique characteristics and underlying reasons for the poorer outcomes in the MSS population is crucial. This study aims to shed light on these aspects, thereby providing invaluable data to guide future drug development.

In colorectal cancer, gene mutations and epigenetic modifications serve as critical but differentially acting molecular mechanisms. Gene mutations involve permanent alterations in the DNA sequence, thereby directly affecting the structure and function of the encoded proteins and playing a pivotal role in disease initiation and progression^[13]^. On the other hand, epigenetic modifications, such as DNA methylation and histone modifications, can influence gene expression without altering the DNA sequence itself and are often reversible ^[14]^. While gene mutations such as KRAS, BRAF, and TP53 are commonly implicated in colorectal cancer^[15]^, epigenetic alterations also contribute to tumor behavior and may serve as therapeutic targets ^[16]^. Both types of molecular changes can work synergistically to influence the tumor microenvironment, tumor progression, and treatment response^[17]^, necessitating their individual consideration in both diagnosis and treatment strategies.

## Materials and Methods

Study Population: Inclusion and Exclusion Criteria

### Inclusion Criteria

1. Patients with a histopathologically confirmed diagnosis of colorectal cancer. 2. Age of 18 years or older. 3. Patients who have undergone genetic testing, microsatellite instability testing, and immunohistochemistry. 4. Comprehensive clinical records are available, including examination results and test results.

### Exclusion Criteria

1. Patients with other forms of malignant tumors. 2. Individuals under the age of 18. 3. Cases with incomplete clinical or pathological data. 4. Patients who have not been subjected to pathological examination or genetic or microsatellite instability testing.

### Data Collection

Data Collection: The data collection process involves gathering comprehensive patient medical records. This includes basic demographic information such as age, gender, and more specific clinical parameters. These parameters encompass tumor differentiation grade, location of the tumor, distant metastasis, TNM staging, and maximum tumor diameter. Additionally, information on nerve invasion and the presence of vascular carcinoma emboli will be collected. Laboratory tests will include tumor markers such as CEA and AFP from peripheral blood, as well as routine blood tests and blood biochemistry. Immunohistochemical assays for markers such as P53, Ki67, and EGFR will also be part of the data set. Last, genetic testing results for MSI, KRAS, BRAF, and other relevant genes will be included.

### Statistical Analysis Methods

Statistical Analysis Methods: The primary statistical analyses will be conducted using RStudio and Excel. The focus of the evaluations will be on chi-square tests to assess the association between categorical variables. When sample sizes are small or expected frequencies are low, Fisher’s exact test will be employed as an alternative to the chi-square test. The approximate Spearman correlation test will be used to examine nonparametric relationships between variables. Visual representations of data distributions and intergroup differences will be illustrated using violin plots and box plots. Mosaic plots will be employed for a graphical overview of relationships between two categorical variables. Statistical significance will be determined at a p value less than 0.05.

## Results

### 1. Baseline Data: Basic information and clinical characteristics of patients

We collected data from 2017 to 2022 for patients in our hospital who underwent curative surgery for colorectal cancer and completed postoperative genetic testing, totaling 325 samples. Due to the relatively low proportions of MSI-L and MSI-H tumors, we tried to balance the number of all three categories during data collection. A table with a total sample size of 325 was created, and the distribution of genetic information is as follows:

**Figure 1.**
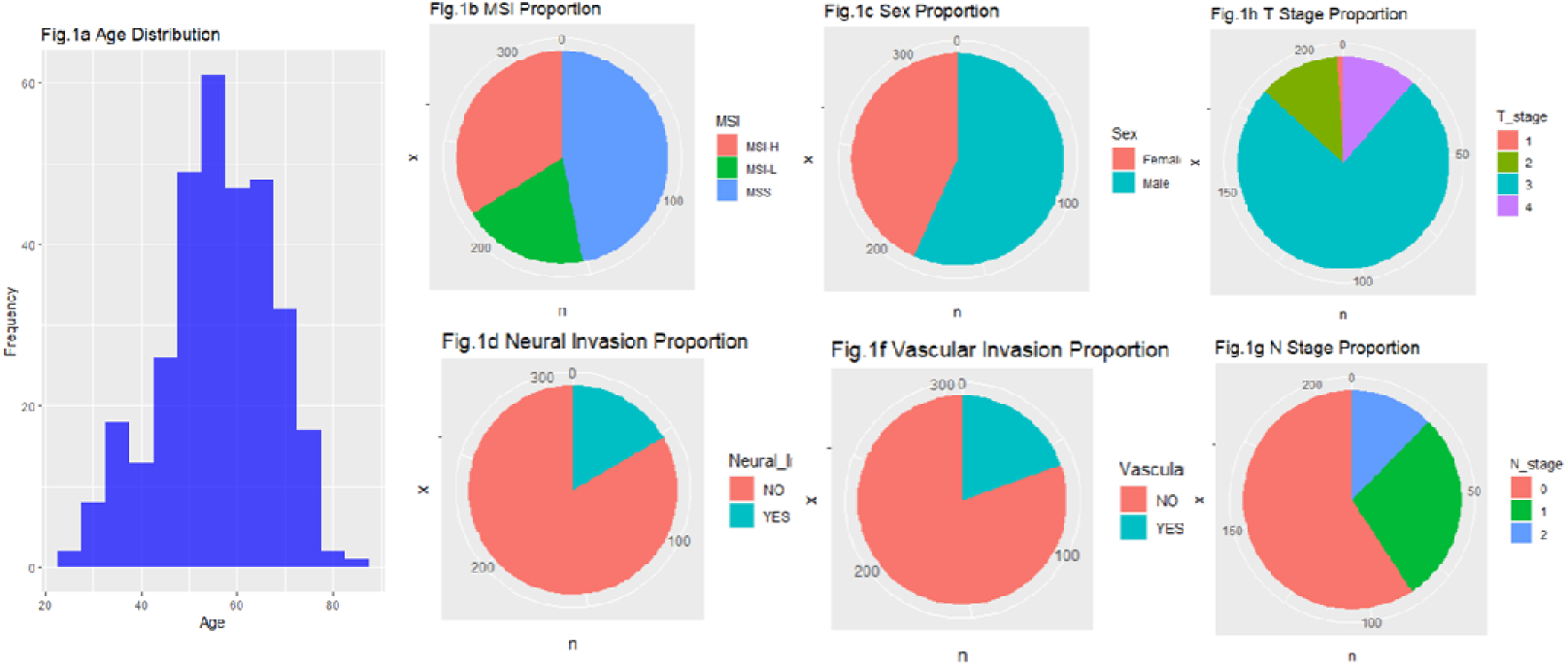
Clinical Baseline Characteristics of the Collected Data Note: Figure 1 illustrates the distribution of various clinical baseline characteristics among the study population, including age, microsatellite instability (MSI), sex, T stage, N stage, perineural invasion, and vascular invasion. The data are presented in various graphical formats for comprehensive interpretation.

### 2. Exploring the Clinical Characteristics of the MSS Group

We divided the samples into three groups, MSS, MSI-H, and MSI-L, and compared them with over 50 clinical indicators (including smoking, drinking, family history, and other exposure histories, as well as routine blood tests, tumor markers, routine biochemical tests, gene test results, pathology results, and immunohistochemistry results) to identify clinical indicators that were significantly different in the MSS group. We selected several meaningful indicators as follows:

**Figure 2.**
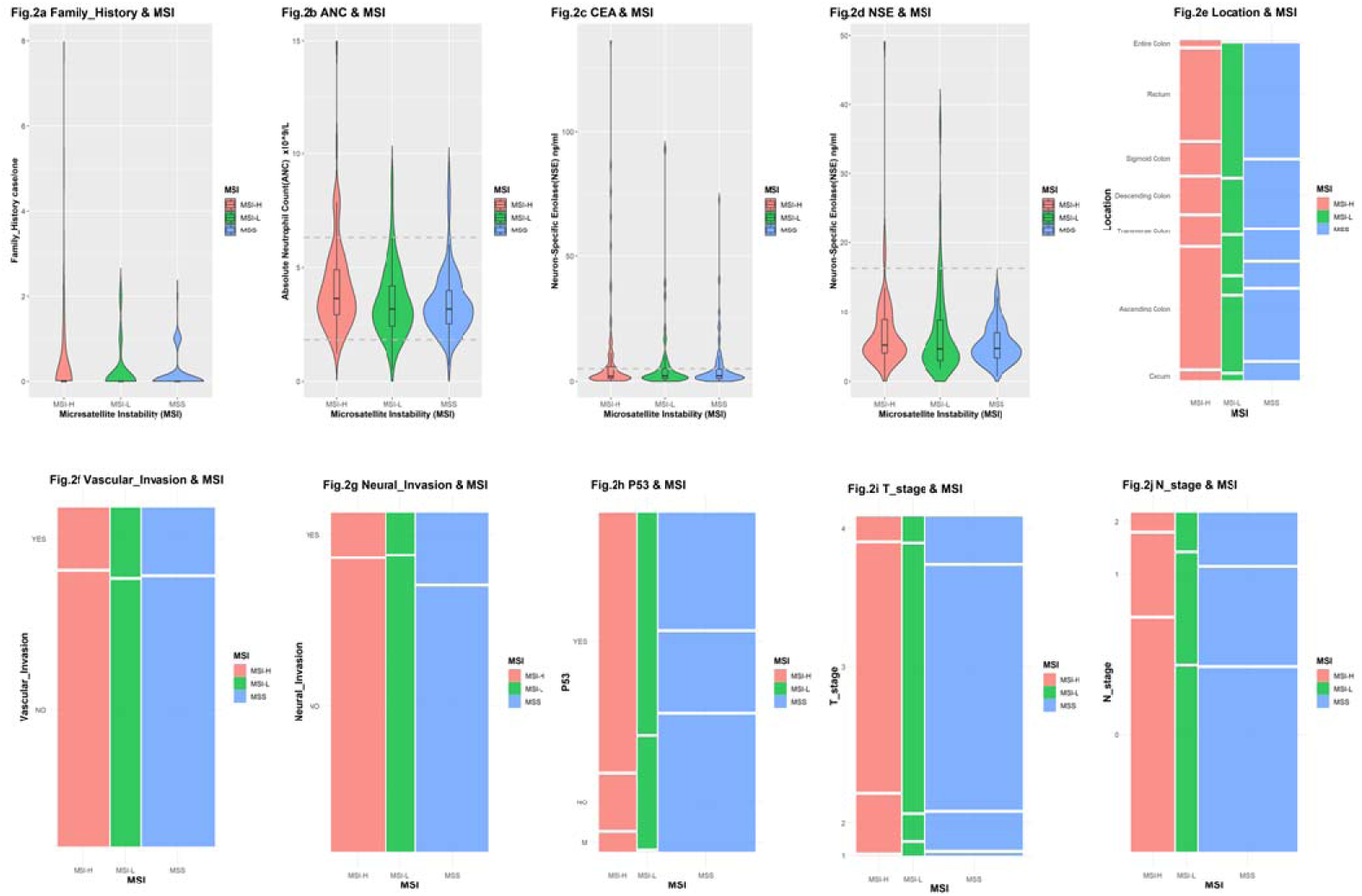
Violin and Mosaic Plots of Clinical Parameters Associated with MSI Subtypes Note: Figure 2 presents violin and mosaic plots for various clinical metrics concerning MSI subtypes. Fig. 2a portrays the relationship between MSI subtypes and the number of family members with cancer history. Fig. 2b elucidates the association between MSI subtypes and neutrophil count. Fig. 2c delineates the correlation between MSI subtypes and peripheral blood CEA levels. Fig. 2d illustrates the relationship between MSI subtypes and peripheral blood NSE levels. Fig. 2e reveals the connection between MSI subtypes and tumor location. Fig. 2 depicts the association between MSI subtypes and postoperative macroscopic pathological specimens indicating vascular invasion by the tumor. Fig. 2g demonstrates the relationship between MSI subtypes and postoperative macroscopic pathological specimens, indicating neural invasion by the tumor. Fig. 2h displays the association between MSI subtypes and postoperative macroscopic pathological specimens featuring P53 mutations (Mut.), abnormal activation (YES), or no expression (NO). Fig. 2i highlights the relationship between MSI subtypes and postoperative macroscopic pathological specimens indicating T stage. Fig. 2j outlines the association between MSI subtypes and postoperative macroscopic pathological specimens indicating N stage. Notably, Fig. 2a, Fig. 2h, and Fig. 2j suggest potential significant differences as indicated in the plots.

**Table 1.** Chip-square test for basic information.

| Varies | Frequency | $\chi^2$ | p value |
| --- | --- | --- | --- |
| Gender | | $\chi^2 = 0.92$ | p value=0.62 |
| Female | 140 |  |  |
| Male | 184 |  |  |
| P53 | | $\chi^2 = 15.06$ | p value=0.004 |
| Mutation | 20 |  |  |
| YES | 36 |  |  |
| NO | 17 |  |  |
| Vascular Invasion | | $\chi^2 = 0.164$ | p value=0.920 |
| YES | 60 |  |  |

**Table 2.** Approximative Spearman Correlation Test for T, N, location Note: Table 2 presents the outcomes of the Approximative Spearman Correlation Test, conducted on the data depicted in Figure 2. For the purpose of this analysis, ordinal variables were assigned as follows: MSI-H, MSI-L, and MSS were marked as 1, 2, and 3, respectively. N stage categories of 0, 1, and 2 were marked as 0, 1, and 2. T stage categories 1, 2, 3, and 4 were marked as 1, 2, 3, and 4, respectively. Tumor locations, including the entire colon, cecum, ascending colon, transverse colon, descending colon, sigmoid colon, and rectum, were sequentially marked as 0, 1, 2, 3, 4, 5, and 6. Hypothesis testing revealed that both N stage and tumor location are significantly correlated with MSI classification (*p < 0.05).

| Varies | Frequency | Z value | p value |
| --- | --- | --- | --- |
| N stage |  | Z = 2.0164, | p value = 0.042 |
| 0 | 125 |  |  |
| 1 | 60 |  |  |
| 2 | 26 |  |  |
| T stage |  | Z = 1.609, | p value= 0.105 |
| 1 | 2 |  |  |
| 2 | 26 |  |  |
| 3 | 159 |  |  |

In this exploration process, some of our data validated previous conclusions:

1. MSI differences are related to tumor location, with MSI-H more commonly seen in the right half of the colon and multiple sites in the colon, while MSS is more commonly seen in the left half of the colon and rectum^[18]^.
2. Family history is more common in MSI, and the difference is more pronounced in cases with multiple family histories (≥3), which previous research has confirmed may be related to Lynch syndrome^[19]^.
3. MSS is somewhat related to abnormal elevation of neutrophils, confirming previous studies showing that MSI-H may be caused by local recurrent inflammation^[20]^.
4. Individual elevations in CEA and NSE were more common in the MSI-H group, although no significant difference was observed in the overall sample.

We also made some new discoveries:

1. MSI tends to have a lower N stage, and the result is statistically significant, which is also an unexpected finding.
2. The mutation rate of P53 was higher in the MSS group, while the proportion of high expression of P53 was significantly higher in the MSI-H group. The difference is statistically significant.
3. Neural invasion is more common in MSS, but the results did not show statistical significance. However, the figures do show some minor differences, which may be related to the small variance and insufficient sample size.

The relationship between P53 and MSS surprised us. P53 was divided into three groups: normal expression, overexpression, and mutation. The results showed that the MSI-H group mainly exhibited P53 overexpression, while the mutation rate of P53 in the MSS group was significantly higher.

### 3. Investigating the Relationship Between MSS and Epigenetic Changes and Gene Mutations

To further investigate whether MSS is associated with the overexpression of other proteins or gene mutations, we continued to search for clues around immunohistochemistry and gene testing results, with the following findings:

**Figure 3.**
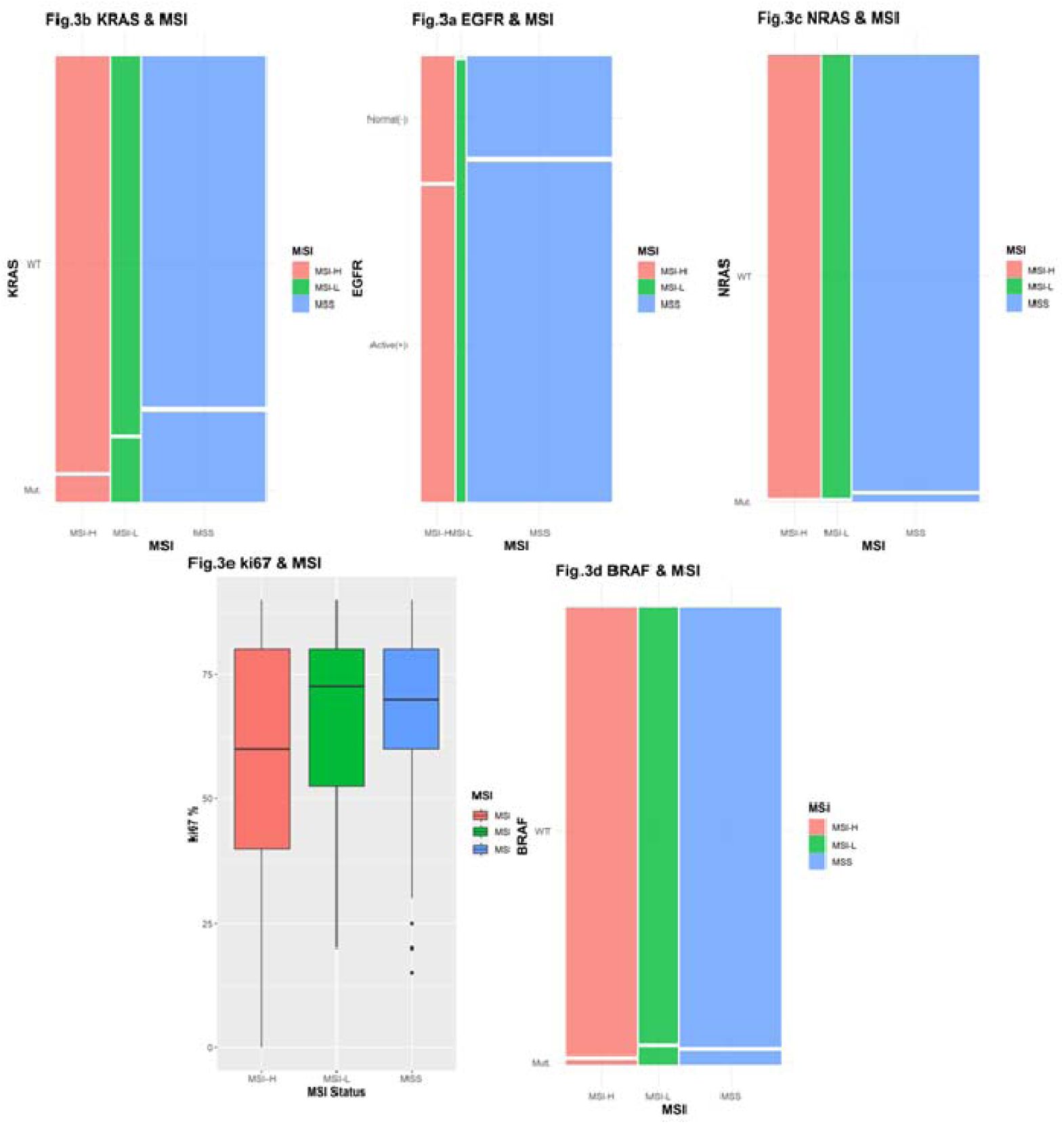
Molecular Characteristics Based on Immunohistochemical and Genetic Data. Note: Figure 3 delves into the molecular attributes beyond P53 utilizing both immunohistochemical and genetic testing data. Fig. 3a illustrates the differences in EGFR protein activation (active [+]/normal [-]) across MSI subtypes. Fig. 3b portrays variations in the mutation status (Mut/WT) of the KRAS gene across MSI subtypes. Fig. 3c displays disparities in the mutation status (Mut/WT) of the NRAS gene among different MSI subtypes. Fig. 3d reveals variations in the mutation status (Mut/WT) of the BRAF gene across MSI subtypes. Fig. 3e exhibits the differences in the percentage of ki67 protein activation expression across MSI subtypes. In the figure, Fig. 3b, Fig. 3c, and Fig. 3d all show that the MSS group has a relatively higher mutation rate compared to the other two groups.

**Table 3.**
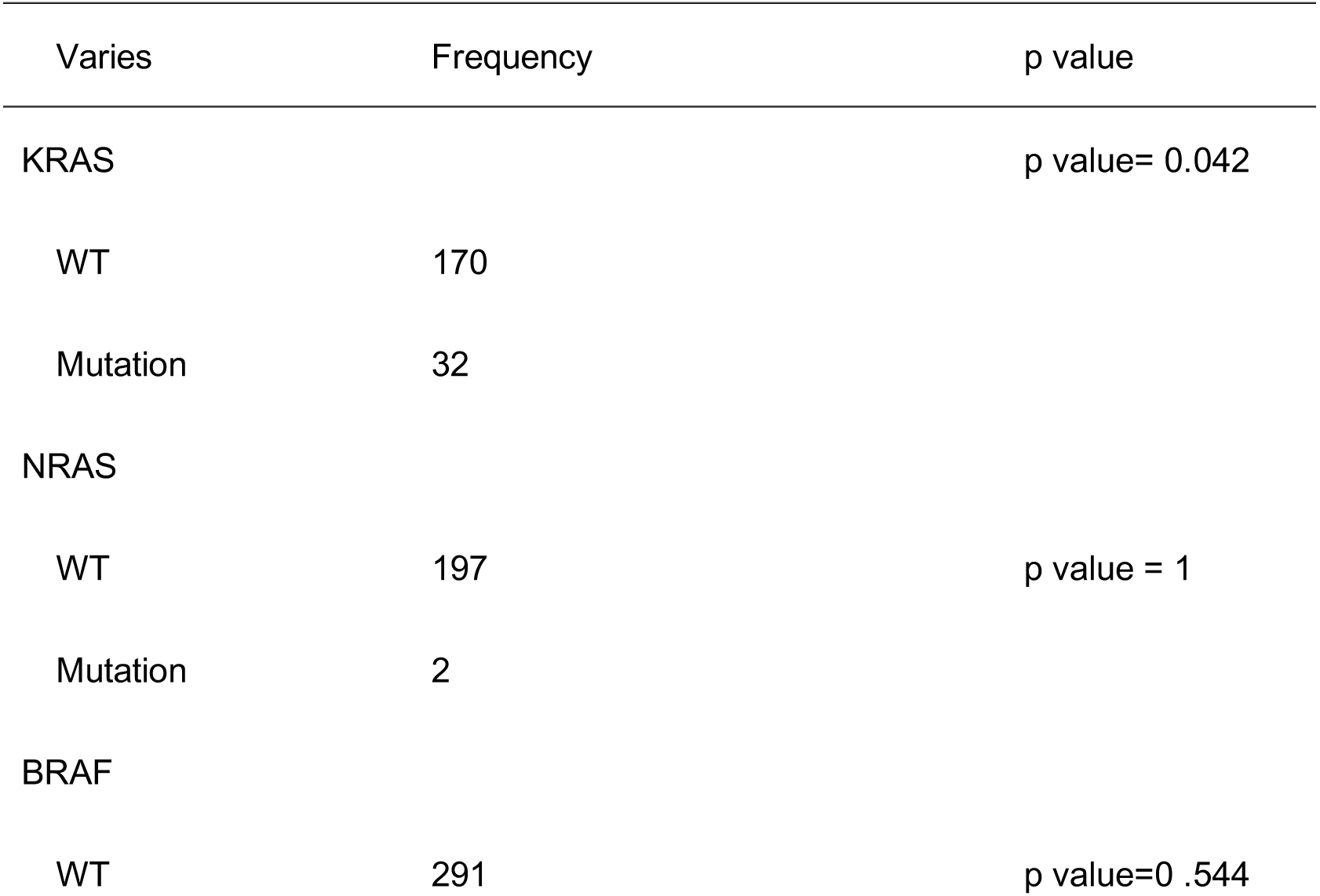
Fisher’s Exact Test for KRAS, NRAS, BRAF, EGFR.

| Varies | Frequency | p value |
| --- | --- | --- |
| KRAS |  | p value= 0.042 |
| WT | 170 |  |
| Mutation | 32 |  |
| NRAS |  |  |
| WT | 197 | p value = 1 |
| Mutation | 2 |  |
| BRAF |  |  |
| WT | 291 | p value=0.544 |

The results show the following: 1. The KRAS gene has a significantly higher mutation rate in the MSS group; 2. The only two NRAS mutation samples were in the MSS group; 3. No significant difference was observed in BRAF mutations among the groups; 4. No significant difference was observed in EGFR among the groups; 5. No significant difference was observed in Ki67 among the groups.

Ki-67 is a protein commonly used as a marker for cell proliferation. It is expressed only during the active phases of the cell cycle, namely, G1, S, G2, and M phases, but not in cells that are in the resting stage (G0 phase). In clinical applications, the Ki-67 index is often measured through immunohistochemical methods to assess the proportion of cells expressing Ki-67 in tumor tissue samples. In this context, we did not observe any significant differences in Ki-67 expression among the different groups^[21]^.

Combining this with previous statistical results on P53, it appears that the MSS group is more likely to have gene mutations, while no significant differences in epigenetic changes such as EGFR and Ki67 were observed between it and the other two groups. Based on this, we suspect that colorectal cancer in the MSS group may be correlated with an overall higher gene mutation rate.

### 4. The overall gene mutation rate was higher in the MSS group for colorectal cancer

To further validate our hypothesis, we summarized the results of immunohistochemistry and genetic testing, categorizing the total sample into groups with gene mutations and those without. The statistical results are as follows:

**Fig. 4.**
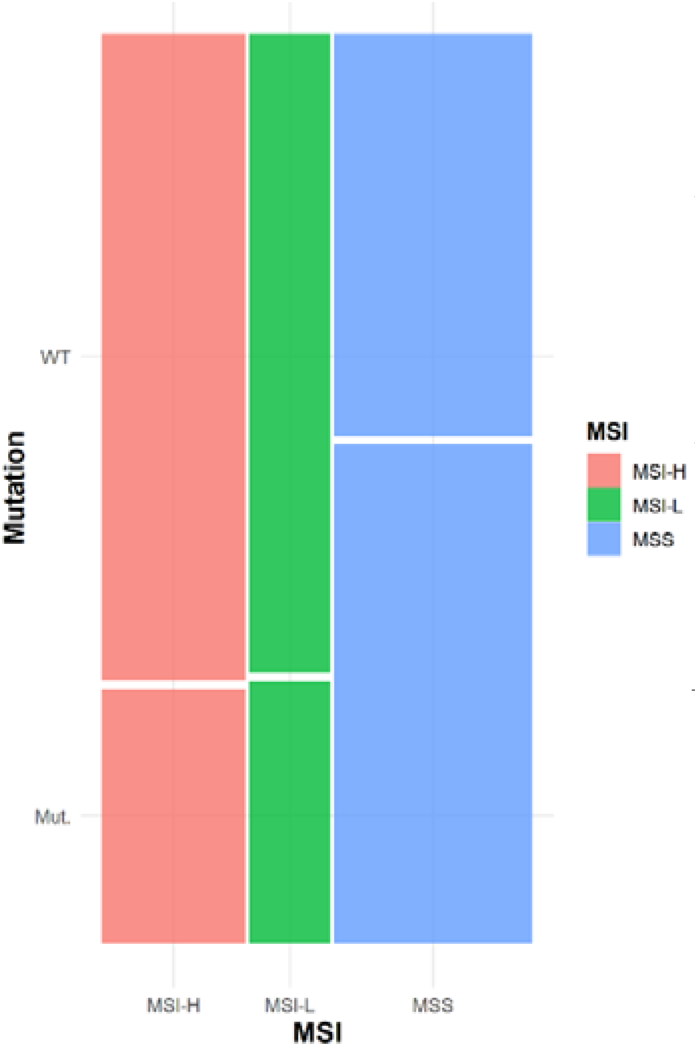
M**utation & MSI** Note: Figure 4 represents a comprehensive analysis of all statistically evaluable data, focusing primarily on results from immunohistochemical assays and gene mutation tests. Samples with identifiable mutations were allocated to the ’Mutated’ category, while those without detectable mutations were classified under the ’Non-Mutated’ category. As illustrated in Figure 4, the MSS subtype shows a significantly higher mutation occurrence rate than the other two MSI subtypes.

**Table 4.**
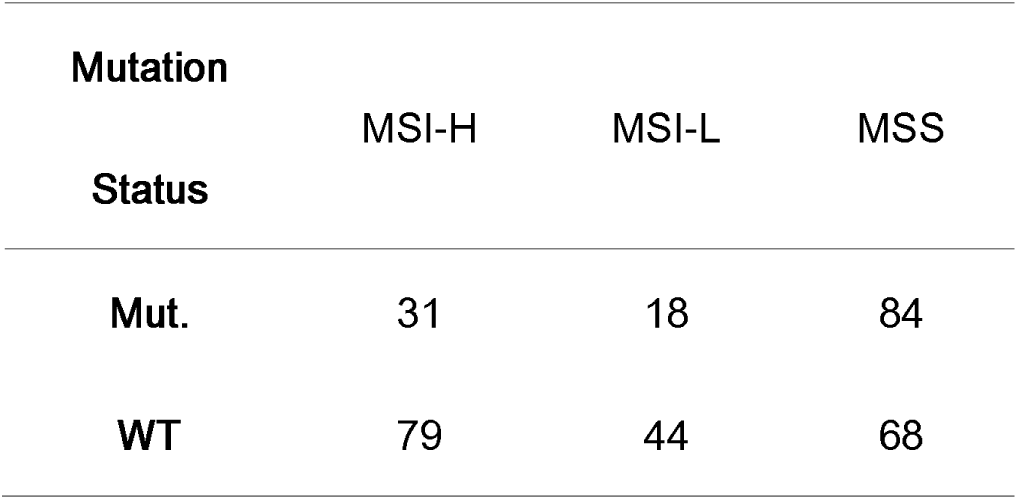
Distribution of MSI Categories by Mutation Status. Note : The table shows the number of occurrences for each combination of Mutation Status (Mut. for Mutated and WT for Wild Type) Note: Table 4 extends the statistical analysis presented in Figure 4. Hypothesis testing was performed to compare the mutation occurrence rates among different MSI subtypes. Notably, the results indicate that the differences in mutation rates are statistically significant (*p < 0.05), particularly showing that the MSS subtype has a higher mutation occurrence rate than the other two subtypes.

The results show that since the p value is far below the conventional significance level of 0.05, we reject the null hypothesis that mutation status and microsatellite instability are independent. This means that there is a significant association between mutation status and microsatellite instability.

### 5. No relationship was found between MSS and environmental exposure

It is well known that genetic mutations are a cause of colorectal cancer rather than a result of it. We wanted to further explore which environmental exposure factors are related to MSS. The results are as follows:

**Figure 5.**
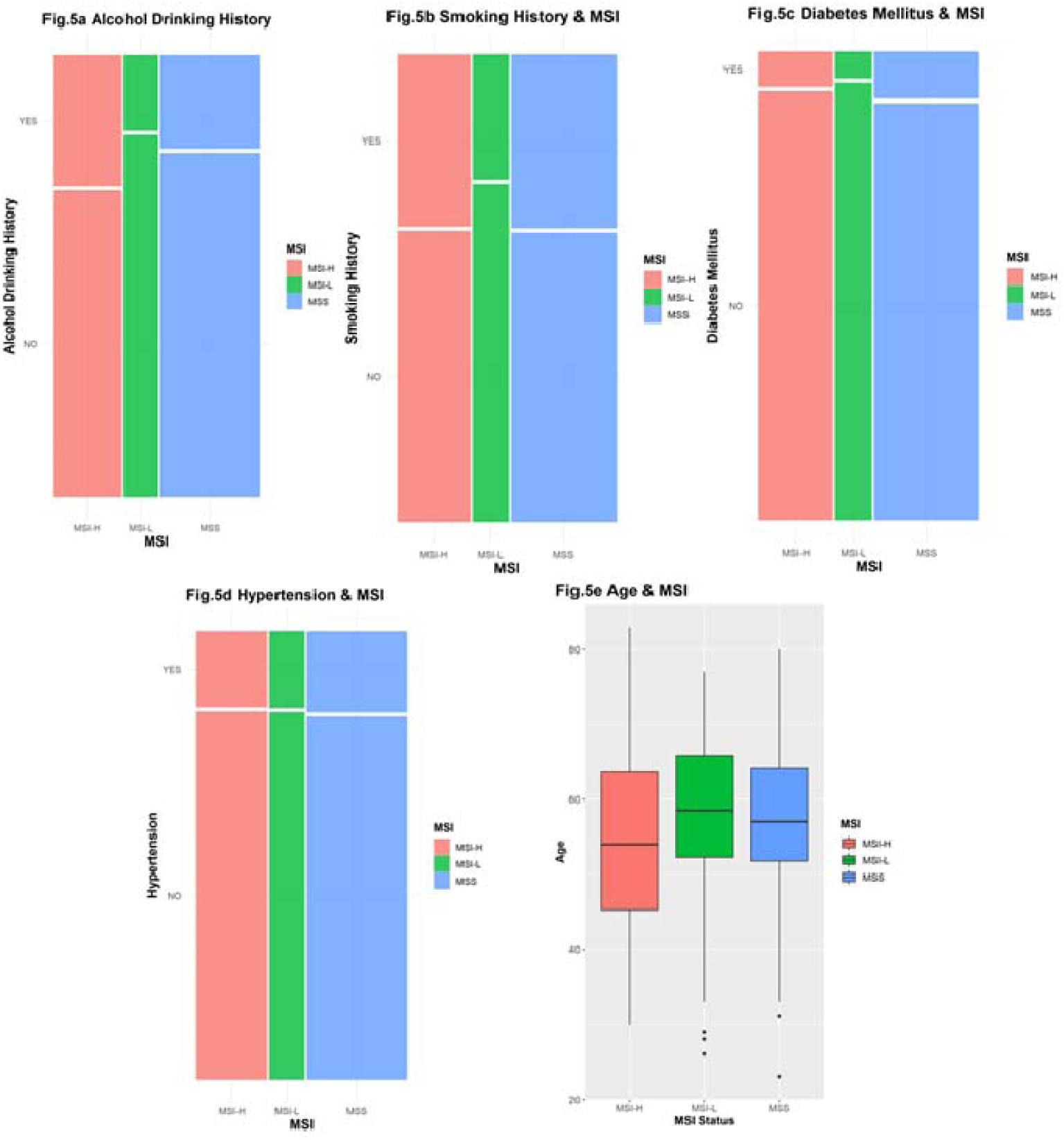
Environmental Factors Potentially Contributing to High Mutation Rates in MSS Subtype. Note: This figure explores the relationship between various environmental factors and the MSI subtypes, specifically focusing on potential causes for the elevated mutation rates observed in the MSS group. Fig. 5a: The correlation between MSI subtype and alcohol consumption. Fig. 5b: Investigates the association between MSI subtype and smoking history. Fig. 5c: Presents data on the relationship between MSI subtype and diabetes. Fig. 5d: Explores the link between MSI subtype and hypertension. Fig. 5e: Analysis of the correlation between MSI subtype and age.

We selected a few representative results and did not find any significant indicators. This suggests that the risk factors leading to MSS genetic mutations may lie in some other more elusive information.

## Discussion

We found that the overall rate of genetic mutations was higher in the MSS group, while the changes in protein expression levels caused by epigenetic changes did not show significant differences among the MSS, MSI-H, and MSS groups. This phenomenon is particularly evident in the immunohistochemical data for P53. This suggests that the poor prognosis of MSS may be related to a higher rate of genetic mutations.

Compared to other studies^[22, 23]^, our research has more comprehensively examined the differences in clinical indicators related to MSS, covering over 50 statistical indicators and then summarizing the results with significant differences. Second, our research focuses more on the uniqueness of MSS, providing informational support for future drug development.

Due to the limitations of the information available, we were unable to identify environmental exposure factors that lead to an increased rate of genetic mutations in the MSS group. Future research should aim to compile as many exposure factors related to the causes of colorectal cancer as possible, such as a preference for a red meat diet.

The aim of our research is to identify distinct characteristics of MSS, suggesting that future drug development should focus more on the field of exonic gene mutations in colorectal cancer. At the same time, efforts should be made in this area to find risk factors leading to poor prognosis. Additionally, to understand the variability and implications of microsatellite instability (MSI) across different subgroups, there is a need for more precise foundational research to quantify the cumulative mutation rates among these subgroups.

## Conclusion

After collecting and organizing data, followed by a series of statistical analyses, we found that the rate of gene mutations in the MSS group with colorectal cancer was significantly higher than that in the MSI-H and MSS groups. This, combined with the relatively poor prognosis observed in the MSS group, suggests a certain relationship between the two. Meanwhile, no significant differences were observed in epigenetic molecular data between the MSI subgroups. We also found that the N stage was lower in the MSI-H group. These findings offer some insights for future drug development and targeted clinical treatments.

## Data Availability

All data produced in the present study are available upon reasonable request to the authors

